# Applications of Artificial Intelligence in clinical decision-making and technical support in Oncology: A Scoping Review protocol

**DOI:** 10.1101/2025.05.14.25327520

**Authors:** Edith Chamoso Alaca, Cyndhi Aránguiz, Javier Bracchiglione, Xavier Bonfill Cosp

## Abstract

**Introduction:** The management of cancer care generates vast amounts of data, collected in the clinical registry; however, the interpretation of these unstandardized and heterogeneous records is often challenging. Artificial intelligence (AI) is emerging as a promising tool in healthcare, particularly in Oncology, where early diagnosis, complex therapies, long term follow-up and the impact on quality of life are paramount. This technology offers significant opportunities to support clinical decision-making and optimize the management of clinical data. Nevertheless, the literature describing how AI is concretely applied to support decision-making and administrative workflow in oncology remains scattered.

**Objective:** The aim is to identify evidence regarding the use of AI in Oncology, focusing on its role in supporting clinical decision-making and technical procedures across the entire cancer care continuum.

**Methods:** This scoping review will analyze primary studies published between 2020 and 2025. Eligibility criteria will be determined based on the PCC framework: Population (cancer patients), Concept (AI), and Context (clinical records). Studies focusing solely on population registries, those lacking identifiable AI use, as well as narrative reviews, will be excluded.

Relevant literature will be identified through a systematic search in MEDLINE, EMBASE and CENTRAL. Additionally, forward citation tracking and review of existing systematic reviews will be conducted. Two independent reviewers will screen articles by titles and abstracts and consequently by full-text, using RAYYAN software. Data extraction will include the following items: cancer type, specific AI technique, the model or tool used, clinical utility, primary recipient (patient or health professional), stage of implementation, and reported outcomes or limitations.

Quality assessment or bias evaluation will not be performed. Findings will be synthesized narratively and, where appropriate, represented graphically.

**Potential Results:** This research is expected to provide a comprehensive synthesis of AI applications in Oncology, highlighting the current role of this technology in enhancing the management of medical records and supporting clinical decision-making in the cancer care continuum. Expected outcomes include evidence of an upward trend in recent publications on the subject, a classification of techniques and models, a catalog of approved AI-based tools, distinctions between patient-oriented and healthcare professional support tools, and the identification of key limitations.

**Conclusion:** The findings of this review will enable clinicians and patients to gain insights into the current landscape of AI in Oncology, identify future research directions, and develop evidence-based strategies for the responsible integration of AI into cancer care. The goal is to position this technology as a supportive tool that complements, rather than replaces, healthcare professionals.

## INTRODUCTION

Cancer remains a major global health burden and one of the leading causes of morbidity and mortality. According to the latest GLOBOCAN report (1), approximately 20 million new cancer cases and 9.7 million related deaths were recorded in 2022. Furthermore, the International Agency for Research on Cancer (IARC) predicts that the annual number of new cancer cases will reach more than 32 million by 2045 (2). The Spanish Society of Medical Oncology (SEOM) estimates that approximately 286,664 new cancer cases were diagnosed in 2024, which represents a 2.6% increase from the previous year, a rising incidence that poses a challenge for healthcare systems (3).

Clinical records are essential for cancer care and research as they provide key information for a comprehensive overview of a patient’s health status and facilitate continuous improvements in medical care (4). However, managing these records effectively is challenging due to the large volume, complexity, diverse sources, lack of standardization, and the limited availability of human resources (5,6).

In this regard, Artificial Intelligence (AI) offers the possibility to optimize the collection, processing, and analysis of these data through advanced techniques, generative and predictive models, and specialized tools designed to enhance clinical decision making in oncology.

AI refers to a branch of computer science dedicated to developing systems that mimic cognitive functions traditionally associated with human intelligence. AI Watch, the European Commission’s knowledge service for monitoring AI, defines it as systems that operate by perceiving their environment through data acquisition, interpreting both structured and unstructured data, reasoning based on this information, and deciding actions to achieve specific goals (7,8).

Key concepts in this domain include techniques (e.g., Machine Learning, Deep Learning, Natural Language Processing), models (i.e., specific implementations of these techniques trained on particular datasets), and tools (applications incorporating AI models to offer specific functionalities) (9). These components are interconnected and essential for the practical implementation of AI systems in a clinical environment (7).

The evolution of this technology is transforming healthcare. It is currently employed in key areas such as direct medical care (including clinical practice and patient self-management), research, data management, and public health with applications ranging from the detection and monitoring of disease outbreaks to promoting health initiatives (10–12). Moreover, the FDA has approved numerous AI-based medical devices, with most being applied to the complex field of cancer diagnostics (13,14).

These algorithms can analyze radiological and pathology images, facilitate early cancer detection, support personalized medicine, predict patient outcomes and treatment responses, enable virtual patient care, and even assist in rehabilitation, among other applications (14–16). Notable examples include AI-assisted mammography screening (18), prostate cancer biopsy analysis (19), radiation therapy planning (20), brain tumour classification from MRI (21), and mortality prediction in emergency oncology admissions (22).

Furthermore, AI has the potential to reduce administrative burdens by extracting valuable information from unstructured electronic health records and clinical notes, converting voice to text, retrieving key data from past medical records, and even detecting prescribing errors in clinical settings (23,24).

By processing vast clinical datasets, AI can identify complex patterns that facilitate medical decision-making across various stages of care, including diagnosis, stratification, prognosis, adverse event prediction, treatment planning support, and clinical follow-up (16,27,28). Despite these promising advancements, AI still faces barriers related to data quality, ethical considerations, and acceptance within clinical settings (12,25,26).

The rapidly increasing number of publications on AI applications in Oncology underscores the need for a structured synthesis to clarify the most actively researched clinical areas, the AI techniques and models employed, and the tools currently approved and in use. Given the imminent integration of AI tools into healthcare settings, this research intends to provide healthcare professionals with a clear overview of the current landscape, identify research gaps, and facilitate the formulation of future research questions. Consequently, this scoping review seeks to explore what are the current clinical applications of AI in supporting decision-making and technical assistance in cancer patient care according to the available scientific evidence.

To address this question, we have defined the following specific objectives:

- to identify techniques, models, and AI-based tools currently employed throughout the cancer care continuum;
- to classify the main areas of AI application by cancer type, direct patient benefit or healthcare professionals, and the stage of care;
- to explore the limitations and challenges associated with the use of AI in this context.

## METHODS

The proposed scoping review will follow the JBI methodology for scoping reviews (29,30) and will be reported according to the Preferred Reporting Items for Systematic Reviews and Meta-Analyses Extension for Scoping Reviews (PRISMA-ScR) guidelines (31).

### Eligibility criteria

We will include primary quantitative and qualitative studies published between January 1, 2020, and May 2025.

Eligibility will be determined based on the following criteria:

#### Population

Studies focusing on cancer patients of all ages, encompassing all forms and stages of cancer.

#### Concept

To be included, a study must incorporate AI. For the purpose of this review, AI is defined as computer systems capable of environmental observation, learning, and task execution with minimal human intervention. This definition covers various AI techniques, models, and tools (7).

#### Context

The context is defined as the application of AI to enhance medical decision-making or provide technical support based on oncology clinical records.

#### Exclusions

We will exclude studies focusing solely on primary care or population-based cancer registries, those in which the specific AI techniques, models, or tools are not clearly identified, and non-empirical formats such as narrative reviews, letters to the editor, conference abstracts, or expert opinions.

### Information sources and search strategy

The search will be conducted until May 9, 2025, across several electronic databases: MEDLINE (via PubMed), EMBASE (via Ovid), and CENTRAL. We will restrict the search to articles published in the last 5 years. Appendix 1 details the search strategies.

Additionally, we will screen reference lists of literature and employ forward and backward citation tracking. Articles included in systematics reviews will also be examined to identify further relevant studies.

### Data management and selection process

We will export search results to the RAYYAN platform for storage, deduplication and screening. Two independent reviewers will screen references by title and abstract, subsequently will screen by full text.

Reasons for exclusion at the full-text stage will be documented and reported. Any discrepancies or uncertainties will be resolved through discussion or by consulting an additional reviewer. A flow diagram, in accordance with PRISMA-ScR guidelines, will visually represent the study selection process.

Consistent with scoping review methodology, no formal quality appraisal or risk of bias assessment will be performed (30).

### Data extraction

We will systematically extract data using a pre-designed data charting developed in Microsoft Excel. This extraction tool will be piloted on a small sample of studies and refined as necessary to address the research objectives. One author will extract all the data, with a second author cross-checking this process. For each included article, key characteristics (e.g., authors, publication year, study design and location) will be recorded. Data items collected are summarized in table 1.

**Table 1.** Data items to be collected from the included studies.

| Item | Operational definition | Categories/Examples |
| --- | --- | --- |
| Type of Clinical Record | Identifies the data sources | Electronic Health Records, Radiology/Imaging Systems, Pathology Records, Genomic/Molecular Databases |
| AI Technique | Specifies the basic methods/ algorithms employed | Machine Learning, Deep Learning, Natural Language Processing |
| AI Model | Describes the concrete implementations | Neural Networks, Large Language Models |
| AI Tool | Lists the specific applications or platforms used (patented, approved or in process). | Specific software names if mentioned |
| Type of Cancer | Categorizes based on affected organ or system | Gynecological, Hematological, Gastrointestinal, etc |
| Direct Recipient | Distinguishes whether the primary beneficiary is the patient or the health professional | Patient, Healthcare professional |
| Application | Differentiates between technical support or medical decision support | Administrative support: voice typing, schedules, manage entering data, etc.<br>Medical decision support |
| Stage of care process | Maps the application across the cancer care continuum | Diagnosis, Staging, Treatment, Follow-up, Prediction of outcomes/toxicity |
| Limitations | Documents barriers or challenges to AI implementation as reported in the studies | As reported in each study |

### Data synthesis

The general features and findings of the included studies will be summarized in tabular form, accompanied by a narrative synthesis outlining key insights.

Data visualization techniques (e.g. bar charts, tree map, bubble map or multilevel donut chart) will be employed to depict trends and classifications within the dataset, thereby mapping the research landscape (32).

## STRENGTHS AND LIMITATIONS

While previous reviews have focused on narrow applications of AI (such as diagnosis or therapeutics), this scoping review aims to provide a more comprehensive synthesis of evidence across all cancer types and clinical settings, including AI tools that directly impact patients. A major strength of this protocol is its adherence to established scoping review guidelines (JBI, PRISMA-ScR), which ensures a robust and reproducible methodology.

However, certain limitations must be considered. As a scoping review, this study will not assess the quality of included articles, since its primary aim is to map the existing literature rather than evaluate methodological rigor. Another limitation is the restriction of the search date to the last 5 years, for which we could potentially omit some older AI applications that may still be relevant. Despite these limitations, the proposed methodology is well suited to address the research question and provide an updated overview of the field, thereby laying the foundation for future research and guiding the responsible implementation of AI into cancer care.

## Data Availability

All data produced in the present work are contained in the manuscript

## ETHICAL CONSIDERATIONS

As this scoping review is based entirely on publicly available secondary data, it does not involve direct human participation or the collection of sensitive, confidential, individually identifiable patient information. The protocol emphasizes adherence to ethical principles, including transparency, respect for intellectual property rights, proper citation, and acknowledgment of original works.

## Appendix 1. Search strategy for each databases

### Search in MEDLINE, via PUBMED

| Search | Query |
| --- | --- |
| #1 | Search: "Neoplasms"[Mesh] OR "Oncology Service, Hospital"[Mesh] OR "Cancer Care Facilities"[Mesh] OR "tumor"[Title] OR "cancer"[Title] OR "neoplasm"[Title] OR "oncolog*"[Title] Filters: in the last 5 years |
| #2 | Search: "Artificial Intelligence"[Mesh] OR "artificial intelligence"[Title/Abstract] OR "machine learn*"[Title/Abstract] OR "natural language proces*"[Title/Abstract] OR "data proces*"[Title/Abstract] OR "deep learn*"[Title/Abstract] OR "NLP"[Title/Abstract] Filters: in the last 5 years |
| #3 | Search: "Routinely Collected Health Data"[MeSH Terms] OR "Registries"[MeSH Terms] OR "Medical Records"[MeSH Terms] OR "Hospital Records"[MeSH Terms] OR "Electronic Health Records"[MeSH Terms] OR "Clinical Decision-Making"[MeSH Terms] OR "Patient Care Management"[MeSH Terms] OR "clinical data"[Title/Abstract] OR "health record*"[Title/Abstract] OR "medical record*"[Title/Abstract] OR "health data"[Title/Abstract] OR "patient care plan*"[Title/Abstract] Filters: in the last 5 years |
| #4 | Search: #1 AND #2 AND #3 Filters: in the last 5 years |

### Search strategy in CENTRAL (Cochrane Library)

|  |  |
| --- | --- |
| #1 | MeSH descriptor: [Neoplasms] explode all trees |
| #2 | MeSH descriptor: [Oncology Service, Hospital] explode all trees |
| #3 | MeSH descriptor: [Cancer Care Facilities] explode all trees |
| #4 | (tumor):ti OR ("Cancer"):ti OR ("neoplasma"):ti OR (oncolog*):ti |
| #5 | #1 OR #2 OR #3 OR #4 |
| #6 | MeSH descriptor: [Artificial Intelligence] explode all trees |
| #7 | ("artificial intelligence"):ti,ab,kw OR ("machine learning"):ti,ab,kw OR ("natural language processing"):ti,ab,kw OR ("natural-language processing"):ti,ab,kw OR ("data processing"):ti,ab,kw |
| #8 | #6 OR #7 |
| #9 | MeSH descriptor: [Routinely Collected Health Data] explode all trees |
| #10 | MeSH descriptor: [Registries] explode all trees |
| #11 | MeSH descriptor: [Medical Records] explode all trees |
| #12 | MeSH descriptor: [Hospital Records] explode all trees |
| #13 | MeSH descriptor: [Electronic Health Records] explode all trees |
| #14 | MeSH descriptor: [Clinical Decision-Making] explode all trees |
| #15 | MeSH descriptor: [Decision Support Systems, Clinical] explode all trees |
| #16 | (health record):ti,ab,kw OR ("medical record file"):ti,ab,kw OR (health data):ti,ab,kw OR (patient care plan*):ti,ab,kw |
| #17 | #9 OR #10 OR #11 OR #12 OR #13 OR #14 OR #15 OR #16 |
| #21 | #5 AND #8 AND #17 with Publication Year from 2020 to 2025, with Cochrane Library publication date Between Jan 2020 and Dec 2025, in Trials |

### Search in EMBASE

| No. | Query |
| --- | --- |
| #1 | 'neoplasm'/exp OR 'cancer center'/exp OR tumor:ti OR cancer:ti OR neoplasm:ti OR oncolog*:ti |
| #2 | 'artificial intelligence'/exp OR 'machine learning'/exp OR 'natural language processing'/exp OR 'artificial intelligence':ti OR 'machine learn*':ti OR 'deep learn*':ti OR 'natural language proces*':ti OR 'data proces*':ti OR nlp:ti |
| #3 | 'medical record'/exp OR 'clinical decision support system'/exp OR 'patient care'/mj OR 'clinical data':ti OR 'health record*':ti OR 'medical record*':ti OR 'health data':ti OR 'patient care plan*':ti |
| #4 | #1 AND #2 AND #3 |
| #5 | #1 AND #2 AND #3 AND [2020-2025]/py |
| #7 | #5 NOT #6 |

